# Sickle Cell Disease Care in Jamaica: Enablers and Barriers to Healthcare Access and Hydroxyurea Utilization

**DOI:** 10.64898/2026.09.16.26363231

**Authors:** Zachary Ramsay, Shelly McFarlane, Onome Braimah, Vanessa Cumming, Jennifer Knight-Madden, Julian Bailey, June Harris, Julia Rowe-Porter, Monika Asnani

## Abstract

Sickle cell disease (SCD) is a significant contributor to the global disease burden, particularly in developing countries where access to care is limited, and coordinated multidisciplinary care is often unavailable. Hydroxyurea is the only readily available disease-modifying-drug therapy in many resource-limited settings, such as Jamaica where it is subsidized through the National Health Fund (NHF). This cross-sectional study assessed barriers to healthcare access and hydroxyurea use in Jamaica among adults with SCD and parents or caregivers of children with SCD from four regional health authorities and a specialized SCD centre. Participants completed an interviewer-assisted questionnaire collecting information on sociodemographics, healthcare access, hydroxyurea utilization and associated barriers. All analyses were exploratory. The study recruited 173 adults (mean age 34.0±11.4 years, 73.6% female, 48.6% urban, 75.1% with secondary education) and 189 parents/caregivers (mean age of parents/caregivers 37.2±10.2 years and children 7.1±4.8 years, 55.0% of children female, 50.3% urban, and 64.3% of parents/caregivers with secondary education). Overall, emergency care was most commonly accessed at public hospitals (87.3%), with the SCU (21.5%) and private hospitals (7.7%) less commonly reported. Routine care was accessed at the SCU by 85.9%, with 28.6% also attending hospital outpatient clinics. Enrolment in the National Health Fund (NHF) drug subsidy programme was reported by 50.9% of adults and 36.0% parents/caregivers, while insurance coverage was reported by 34.7% and 31.8%, respectively. Common healthcare access barriers included not being seen quickly enough when in pain (77.8% adults), worry or fear (47.2% overall), long wait times (39.2% overall), frustration or anger (29.3% overall), and high transportation costs (22.4% overall). Hydroxyurea awareness was reported by 62.4% adults and 67.6% parents/caregivers, while 45.0% and 55.0% reported receiving hydroxyurea education, respectively. Awareness and education were both higher among adult females and parents/caregivers of older children, while awareness was also greater among tertiary-educated adults. Overall, 30.2% of adults and 39.4% of children reported initiating hydroxyurea (p=0.18). Among patients advised to take hydroxyurea, 68.4% adults and 77.1% children initiated (p=0.18), and 50.0% adults and 73.4% children (p=0.001) were currently using it. Among adults, lower initiation was associated with reporting insufficient knowledge about hydroxyurea (odds ratio, OR=0.1), disinterest in another medication (OR=0.1), and having neither NHF coverage nor other health insurance (OR=0.1). Among parents/caregivers, initiation and current use were lower among those who were disinterested in another medication (OR=0.1 and 0.02, respectively) and concerned about side effects (both OR=0.2). Poor adherence was most commonly attributed to forgetting to take the medication. Adherence was lower among adults who reported inadequate social support and healthcare being too expensive, including co-pay. Among children, adherence was greater among females, younger children and those with tertiary-educated parents. Findings suggest hydroxyurea awareness and education are inadequate, and initiation and continued use remain suboptimal, highlighting the need for improved education strategies and NHF enrolment. Strategies should be tailored to better engage males, parents and patients of different age groups, and those with lower educational attainment. Greater support is needed to address mental health and transportation challenges, alongside protocols aimed at reducing wait times.

## Introduction

### Access to care in sickle cell disease

Sickle cell disease (SCD) is associated with a lifetime of medical and socio-behavioural complications that require coordination of care from multidisciplinary teams including physicians trained in SCD management, psychologists, social workers, and pain specialists [1]. Unfortunately, coordinated multidisciplinary care is rarely a reality. Inadequate health care access may contribute to increased acute care utilization, disjointed care delivery, and earlier mortality for many SCD patients [2–4]. Access to care for SCD, especially among adults, has remained challenging globally. In some settings care for children may be better developed, but continuous and comprehensive care services for adults have remained disjointed [5]. Improvements in access to quality care for all patients with SCD are anticipated to improve patients’ quality of life and result in greater cost-effectiveness as compared with episodic, accident and emergency department-based care [6]. Patients with SCD treated at centres able to provide routine care, including health maintenance visits, use emergency and inpatient facilities less frequently, have decreased health care costs overall, and are more likely to be prescribed hydroxyurea [7–10]. Various barriers to adequate care that have been identified include inadequate healthcare provider knowledge, limited patient/caregiver knowledge, stigma and implicit bias surrounding SCD management, frequent healthcare utilization with associated poor provider attitudes to provision of care, and inadequate financial resources [11].

### Hydroxyurea use in sickle cell disease

In individuals living with SCD, higher levels of fetal haemoglobin (HbF) have been found to be protective as HbF inhibits the polymerization of abnormal hemoglobin S (HbS) and sickling [12]. Hydroxyurea is the first disease-modifying drug approved for the treatment of SCD and works mainly by increasing HbF levels, as well as improving many other pathophysiological mechanisms underlying SCD. The clinical effects are evident especially in reducing recurrent severe acute pain events, acute chest syndrome, and hospitalizations, and improving patients’ quality of life [13]. These effects are seen in both children and adults, and in fact, many centres now offer hydroxyurea to children as young as 9 months of age as standard routine care [14].

Despite this clear evidence of benefit and efficacy, the uptake of hydroxyurea and its use remains suboptimal globally, particularly in settings such as Sub-Saharan Africa where the prevalence of SCD is high [15, 16]. There is hesitancy on the part of providers, patients, and families to initiate hydroxyurea therapy and poor adherence when it is begun. Prior studies have explored this issue and found possible explanations to include low awareness of its benefits among both clinicians and patients, perceived risks associated with its use, anticipated burden of increased visits for monitoring, poor availability, or financial burden, among others [16–18].

### Management of sickle cell disease in Jamaica

For many years, the Sickle Cell Unit (SCU) was the only dedicated provider of care for persons living with SCD in Jamaica providing specialized routine and emergency care from its clinic at The University of the West Indies (UWI) [19, 20]. SCD was added to Jamaica’s 2013-2018 National Strategic Plan for Non-Communicable Diseases thus affording it greater visibility and a structured management approach nationally. The public healthcare system in Jamaica is managed by the Ministry of Health and Wellness (MoHW), which decentralizes its operations through four Regional Health Authorities (RHAs). The Sickle Cell Technical Working Group (SCTWG) of the Ministry of Health & Wellness (MOHW), Jamaica was re-activated in 2014 since then training in SCD management has been provided to all regional health authorities (RHA) so that, increasingly, health care providers in all parishes are capable of providing care for patients locally. This training is supported by the adoption of SCU’s Clinical Care Guidelines to Jamaica’s national NCD management protocols (citations are needed for both guidelines and protocols). Provision of care through the 4 RHAs is managed by the regional SCD coordinators, aided by the parish coordinators.

### Hydroxyurea and SCD in Jamaica

The birth incidence of SCD in Jamaica at 1 in 154 live births is the highest in the Caribbean region, which itself remains second only to Sub-Saharan Africa [21]. In the absence of a national SCD registry, it is estimated that 18,000-19,000 persons are living with the disease in the island. Jamaica also has a decades-long history of research and expert clinical care for SCD, yet to date, few studies have explored the use of hydroxyurea in this population. Hydroxyurea has been offered to patients at the SCU for over 20 years. The SCU Clinical Care Guidelines, first published in 2008 and revised in 2015, clearly state the criteria for initiating hydroxyurea in children and adults [22]. The drug is widely distributed within the public health care system and it was added to the government’s National Health Fund (NHF) drug subsidy programme in 2015 for better affordability. Despite these efforts to improve awareness, availability and affordability, less than 10% of the SCU population were taking the drug in 2023 at the inception of this study based on a routine review of internal service data. The use of hydroxyurea for SCD remains limited in the rest of the island. Data from the NHF shows that since the addition of the drug to its formulary in 2015, there have been only 292 claimants island wide up to 2024, with a total of about 1438 claims over the 1-year period prior. The majority of the claims have been concentrated in the Kingston and St Andrew area where the SCU is located.

#### Study aims and objectives

This project sought to conduct an assessment of general access to care for an at-risk population with SCD in Jamaica. Many of the factors that impact general access to care are expected to also impact the uptake of hydroxyurea for these patients as well. The study also aimed to examine specifically the barriers and enablers of hydroxyurea use in Jamaica.

Specifically, the study sought to:

1. Determine where people living with SCD access care and their perceived barriers to receiving care
2. Assess the awareness, education, utilization, and barriers to use of hydroxyurea among patients and caregivers of patients with SCD

## Materials and methods

A cross-sectional quantitative survey study was conducted in Jamaica and recruited adults (ages 18 years or older) living with SCD and immediate caregivers of children (aged less than 18 years) living with SCD. Convenience and snowball sampling was used to recruit participants from four regional health authorities across Jamaica and participants from the SCU at the University of the West Indies (UWI), Kingston. The SCTWG, within the MOHW, Jamaica, has SCD coordinators in each regional health authority who assisted in the recruitment from each region. The study protocol was developed through discussions within the SCTWG which also has representation from patients’ peer network. The study coordinators thereafter administered the surveys once informed consent was received. Surveys were completed either remotely via Zoom or WhatsApp platforms or face-to-face as preferred by the participants. Participants were not recruited if acutely unwell. Data were collected between May 2023 and August 2024. All survey data were stored in a REDCap database, which is an open-source online platform that is secured and hosted by the Mona Information Technology Services (MITS) of the UWI. Ethics approval for the study was received from both the UWI’s Mona Campus Research Ethics Committee (Reference #: CREC-MN.008, 2022/2023) and the MOHW’s Advisory Panel on Ethics and Medico-Legal Affairs (Reference #: 2022/29).

### Measurement Tools

The questionnaire surveys were adapted from the Sickle Cell Disease Implementation Consortium (SCDIC) project which is a multicentre study conducted in the United States [23]. Permission was granted by the SCDIC study administrators to use the tools as needed.

Socio-demographic information included age, sex, education level, employment status, health insurance, and household income. Clinical information was sought from patients and parents/caregivers to understand the severity of the illness experienced. Access to care and various potential barriers to accessing care such as transportation, insurance, provider knowledge and attitudes, health facilities barriers, social support, and individual-specific barriers were examined.

Participants then reported on their awareness and knowledge about hydroxyurea, if they had been recommended to take it, and their usage of and adherence to hydroxyurea. The SCDIC 20-item ‘Barriers to hydroxyurea use checklist’ (response levels: Yes/ No/ Don’t know) was then administered.

### Statistical Methods

Statistical significance was set at p<0.05 and all analyses were done in Stata 16. Among the main variables included in this analysis there was minimal and inconsequential missing data of less than 3% [24, 25]. Sociodemographic and clinical factors, as well as barriers to healthcare access and hydroxyurea use were described as means with standard deviations, medians with interquartile ranges, or cross tabulations with frequencies and percentages, where appropriate. A housing crowding index was calculated by dividing the total number of persons in the household by the total number of rooms used for sleeping [26]. Genotype was collapsed into severe and mild, based on the presence of SS or SB-thalassemia-0 and SC or SB-thalassemia+, respectively [27].

All inferential analyses were conducted separately among the adult patient and the parent or caregiver subsets of the sample. Pearson’s chi-squared and Wilcoxon rank-sum tests were used to assess for differences in hydroxyurea awareness and education, based on sociodemographic characteristics. Among participants currently taking hydroxyurea, Pearson’s chi-squared and Wilcoxon rank-sum tests were used to identify factors associated with hydroxyurea adherence. Given the exploratory nature of these analyses, multiple p-value testing corrections were not performed, and therefore the associations should be interpreted with caution.

Among patients who were advised to take hydroxyurea, univariable logistic regression models were used to identify associated clinical and sociodemographic factors, as well as hydroxyurea-specific barriers, associated with initiation of the drug. A sensitivity analysis was conducted using current hydroxyurea use as the outcome to identify the factors associated with continuation of hydroxyurea. To reduce the imbalance of the categories, adherence was collapsed into two categories, “always” (adherent) and “sometimes or usually” (non-adherent). To reduce the risk of false-positive findings due to multiple comparisons, p-values from the logistic regression analyses were adjusted using the Benjamini-Hochberg false discovery rate (FDR) method [28]. However, both unadjusted and FDR-adjusted p-values are presented to allow transparent interpretation of the exploratory analyses.

For the descriptive analyses, responses of “unsure”, “do not know” or an intentional non-response were kept and analyzed in their original format as these responses are considered valid and meaningful. However, when performing inferential analyses using the hydroxyurea-specific barriers, responses of “don’t know” were handled as missing data as these responses were sparse and resulted in highly imbalanced categories that could not be meaningfully analyzed. All analyses are intended to be interpreted as an observational exploration of barriers to hydroxyurea use, and not as causal inferential relationships.

## Results

There were 362 participants recruited to the study, including 173 adult patients and 189 parents/caregivers of children with SCD. The mean age of the adult participants was 34.0±11.4 years, 128 (73.6%) were female, and 84 (48.6%) were living in urban areas. The mean age of the children was 7.1±4.8 years, 104 (55%) were female, and 95 (50.3%) lived in urban areas. The main parents/caregivers recruited were mothers (n=151; 79.9%) and fathers (n=13; 6.9%). The mean age of the parents/caregivers was 37.2±10.2 years. Almost all children (n=162, 86.2%) lived with one or both parents/caregivers whereas 26.4% of adults lived with a consort and 12.8% lived alone.

### Sociodemographics of the study sample

Table 1 describes the main sociodemographics of the two groups. For adults and parents/caregivers respectively, the mean crowding index was 1.1±0.7 and 1.8±1.2; 61.3% and 38.4% had never married; and 12.1% and 27% were married. Three-quarter (75.1%) of adults and 64.3% of parents/caregivers had secondary education with 18.5% and 32.9% of respective groups having tertiary education. Over two-thirds (67.2%) of parents/caregivers and 56.1% of adult patients were employed full-time.

**Table 1:** Sociodemographic characteristics of the study participants.

|  | **Parent/Caregiver** | **Adult patients** |
| --- | --- | --- |
| Crowding index (mean ± standard deviation) | 1.8±1.2 | 1.1±0.7 |
| Marital status -n, % |  |  |
| Never married | 71 (38.4%) | 106 (61.3%) |
| Married | 50 (27.0%) | 21 (12.1%) |
| Separated or divorced | 6 (3.2%) | 8 (4.6%) |
| Common-law | 50 (27.0%) | 27 (15.6%) |
| Visiting relationship | 8 (4.3%) | 11 (6.4%) |
| Total household monthly income - n (%) |  |  |
| Less than 27,000 | 13 (6.9%) | 14 (8.1%) |
| 27,001-47,999 | 27 (14.3%) | 15 (8.7%) |
| 48,000-99,999 | 19 (10.1%) | 13 (7.5%) |
| 100,000-249,000 | 28 (14.8%) | 24 (13.9%) |
| More than 250,000 | 15 (7.9%) | 19 (11.0%) |
| Unsure | 50 (26.4%) | 52 (30.1%) |
| No response | 37(19.6%) | 36 (20.8%) |
| Employment status - n (%) |  |  |
| Full time (30 or more hours per week) | 127 (67.2%) | 97 (56.1%) |
| Part time (29 or less hours per week) | 16 (8.5%) | 25 (14.5%) |
| Unemployed and looking | 21 (11.1%) | 16 (9.3%) |
| Unemployed and not looking | 21 (11.1%) | 19 (11.0%) |
| Student | 4 (2.1%) | 16 (9.3%) |
| Education (highest level achieved) - n (%) |  |  |
| Primary | 3 (1.7%) | 12 (6.4%) |
| Secondary or vocational | 113 (64.3%) | 142 (75.1%) |
| Tertiary | 57 (32.9%) | 35 (18.5%) |

### Sickle cell disease history

Among both adults and children, the most common genotype was SS disease (adults - SS: n=128, 73.6%; SC: n=34, 19.5%; Sβ+Thalassemia: n=8, 4.6%; Sβ0-Thalassemia: n=4, 2.3%; and children - SS: n=154, 81.9%; SC: n=19, 10.1%; Sβ0-Thalassemia: n=8, 4.3%; Sβ+Thalassemia: n=6, 3.2%) and therefore, 160 children (85.1%) and 132 (76.3%) adults had a severe genotype. More children were diagnosed at birth (n=138, 73.0%) compared to adults (n=51, 29.5%). Five parents/caregivers reported also having SCD (2.7%) and a separate five were unaware of their status (2.7%).

Table 2 provides details of SCD related complications and complementary therapies being utilized. Half the adults (50%) and 66.7% of children had no acute pains in the past year but 13.4% adults and 6.9% of children had 5 and more acute pain crisis in the year prior. The commonest reported lifetime SCD complications for adults included ‘joint issues’ including avascular necrosis of the hip (n=129, 74.6%), acute chest syndrome (n=86, 49.7%), and leg ulcers (n=29, 16.8%). For children, the commonest complications reported were acute chest syndrome (n=97, 51.3%), and joint issues (n-94, 49.7%). A small subset (n=7, 3.5%) of the children had a history of having had a stroke. Almost one-third (32.9%) of adults and 32.5% of children were hospitalized once or more annually with 13.8% adults and 17% of children never been hospitalized.

**Table 2:**
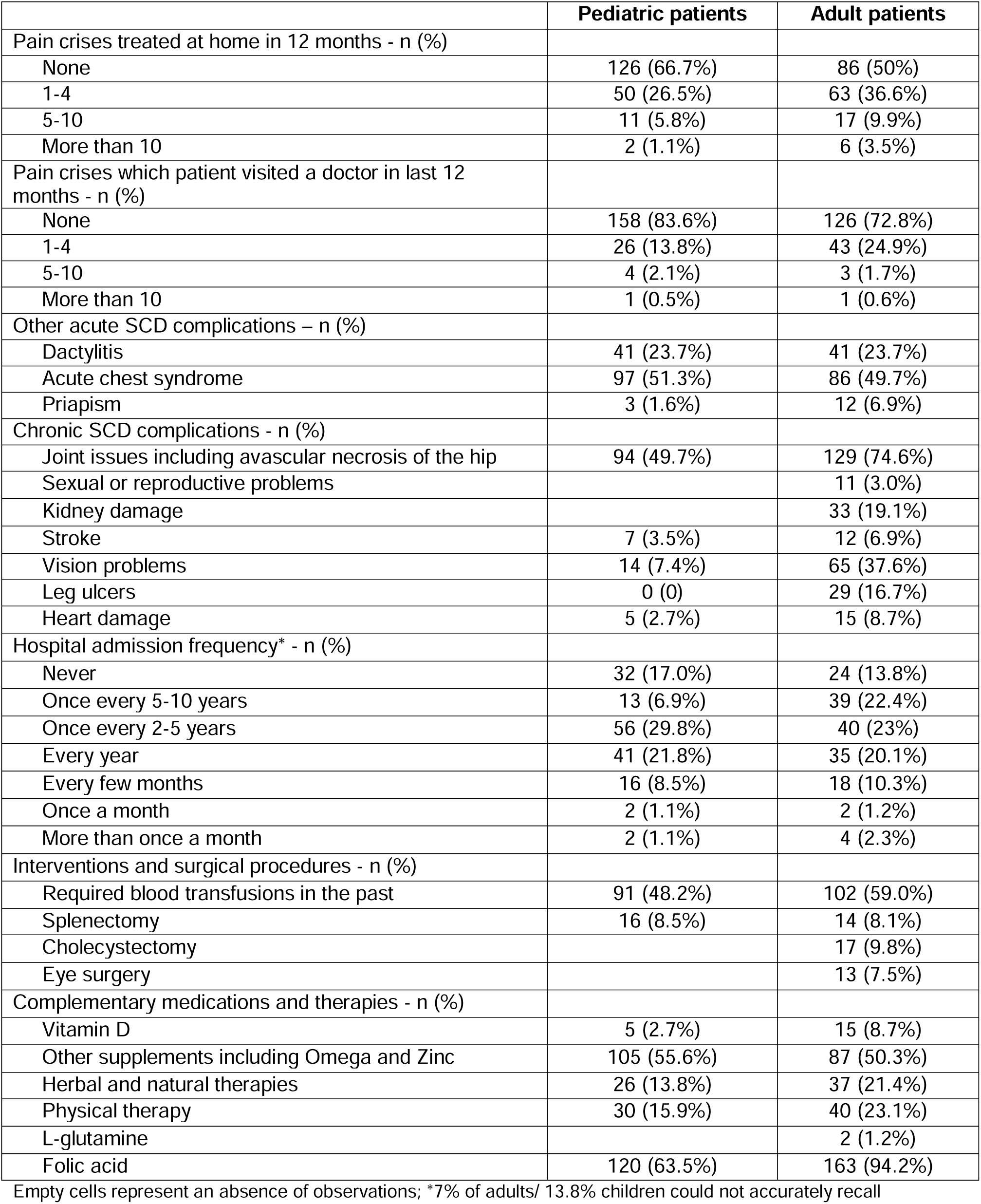
Sickle cell disease complications and interventions reported during lifetime by Jamaicans with sickle cell disease and their caretakers.

|  | Pediatric patients | Adult patients |
| --- | --- | --- |
| Pain crises treated at home in 12 months - n (%) |  |  |
| None | 126 (66.7%) | 86 (50%) |
| 1-4 | 50 (26.5%) | 63 (36.6%) |
| 5-10 | 11 (5.8%) | 17 (9.9%) |
| More than 10 | 2 (1.1%) | 6 (3.5%) |
| Pain crises which patient visited a doctor in last 12 months - n (%) |  |  |
| None | 158 (83.6%) | 126 (72.8%) |
| 1-4 | 26 (13.8%) | 43 (24.9%) |
| 5-10 | 4 (2.1%) | 3 (1.7%) |
| More than 10 | 1 (0.5%) | 1 (0.6%) |
| Other acute SCD complications – n (%) |  |  |
| Dactylitis | 41 (23.7%) | 41 (23.7%) |
| Acute chest syndrome | 97 (51.3%) | 86 (49.7%) |
| Priapism | 3 (1.6%) | 12 (6.9%) |
| Chronic SCD complications - n (%) |  |  |
| Joint issues including avascular necrosis of the hip | 94 (49.7%) | 129 (74.6%) |
| Sexual or reproductive problems |  | 11 (3.0%) |
| Kidney damage |  | 33 (19.1%) |
| Stroke | 7 (3.5%) | 12 (6.9%) |
| Vision problems | 14 (7.4%) | 65 (37.6%) |
| Leg ulcers | 0 (0) | 29 (16.7%) |
| Heart damage | 5 (2.7%) | 15 (8.7%) |
| Hospital admission frequency* - n (%) |  |  |
| Never | 32 (17.0%) | 24 (13.8%) |
| Once every 5-10 years | 13 (6.9%) | 39 (22.4%) |
| Once every 2-5 years | 56 (29.8%) | 40 (23%) |
| Every year | 41 (21.8%) | 35 (20.1%) |
| Every few months | 16 (8.5%) | 18 (10.3%) |
| Once a month | 2 (1.1%) | 2 (1.2%) |
| More than once a month | 2 (1.1%) | 4 (2.3%) |
| Interventions and surgical procedures - n (%) |  |  |
| Required blood transfusions in the past | 91 (48.2%) | 102 (59.0%) |
| Splenectomy | 16 (8.5%) | 14 (8.1%) |
| Cholecystectomy |  | 17 (9.8%) |
| Eye surgery |  | 13 (7.5%) |
| Complementary medications and therapies - n (%) |  |  |
| Vitamin D | 5 (2.7%) | 15 (8.7%) |
| Other supplements including Omega and Zinc | 105 (55.6%) | 87 (50.3%) |
| Herbal and natural therapies | 26 (13.8%) | 37 (21.4%) |
| Physical therapy | 30 (15.9%) | 40 (23.1%) |
| L-glutamine |  | 2 (1.2%) |
| Folic acid | 120 (63.5%) | 163 (94.2%) |

Empty cells represent an absence of observations; \*7% of adults/ 13.8% children could not accurately recall

Self-reported mean scores of overall health were 5.7±1.0 and 6.3±1.0 (possible range: 1 to 7) among adults and children (p<0.001), respectively. Self-reported emotional wellbeing mean scores were 4.3±2.0 and 4.1±2.3 (possible range: 1 to 7) among adults and children (p=0.83), respectively. Higher scores imply better health for both items. Over one-third (37.4%) of adults and 22.9% of children reportedly required support from someone for their daily activities as a result of their SCD. Complementary therapies were reported among both cohorts including folic acid (283, 78.2%), physiotherapy (n=70, 19.3%), herbal therapies (n=63, 17.4%), and Vitamin D supplements (n=20, 5.5%).

### Health care access and barriers

Reported sites of care for emergencies (non-mutually exclusive) included public hospitals among more than 80% of both cohorts (adult: n= 144, 83.2% and children: n=172, 91.0%), while the SCU (adult: 50, 28.9%, and children: 28, 14.8%) and private hospitals (adult: n=24, 13.9% and children: n=4, 2.1%) were less commonly reported. Routine health maintenance care was accessed (non-mutually exclusive) at the SCU by more than three-quarters of both groups (adult n=136, 78.6% and children n=175, 92.6%); with 32% adults and 25.5% children also attending a hospital out-patient clinic for routine visits.

Registration with the National Health Fund (NHF) drug subsidy programme was reported for 68 (36.0%) children and 88 (50.9%) adults. Private insurance through employers was available to 28.3% adults and 26.5% children; and self-insurance was accessed by 6.4% adults and 5.3% children. Twenty-seven adults and 25 children reported having a combination of either different types of insurance, or insurance and NHF coverage. Prescription medications were reported as taken in the last 12 months by 153 children (81.4%) and 156 adults (90.7%). Among these, 41.8% of the parents/caregivers and 52.6% of the adults worried about the cost of medication.

Barriers to healthcare access are summarized in Table 3, and the full table is listed in Supplementary Table S1. The five most common barriers were: not being seen quickly enough when in pain among adults (77.8%); lack of insurance among both parents/caregivers and adults (51.9% and 38.2% respectively), worry or fear (51.3% parents/caregivers and 42.8% adults) and frustration or anger related to their disease (34.7% adult and 24.3% caregivers); long waits at the healthcare facility (45.1% adults and 33.9% caregivers) and transportation costs being too high (19.1% adult and 25.4% caregivers). Adults were more likely to report feeling isolated, not finding an expert provider, inadequate communication with providers, stigma, and frustrations related to their disease as being significant barriers.

**Table 3:**
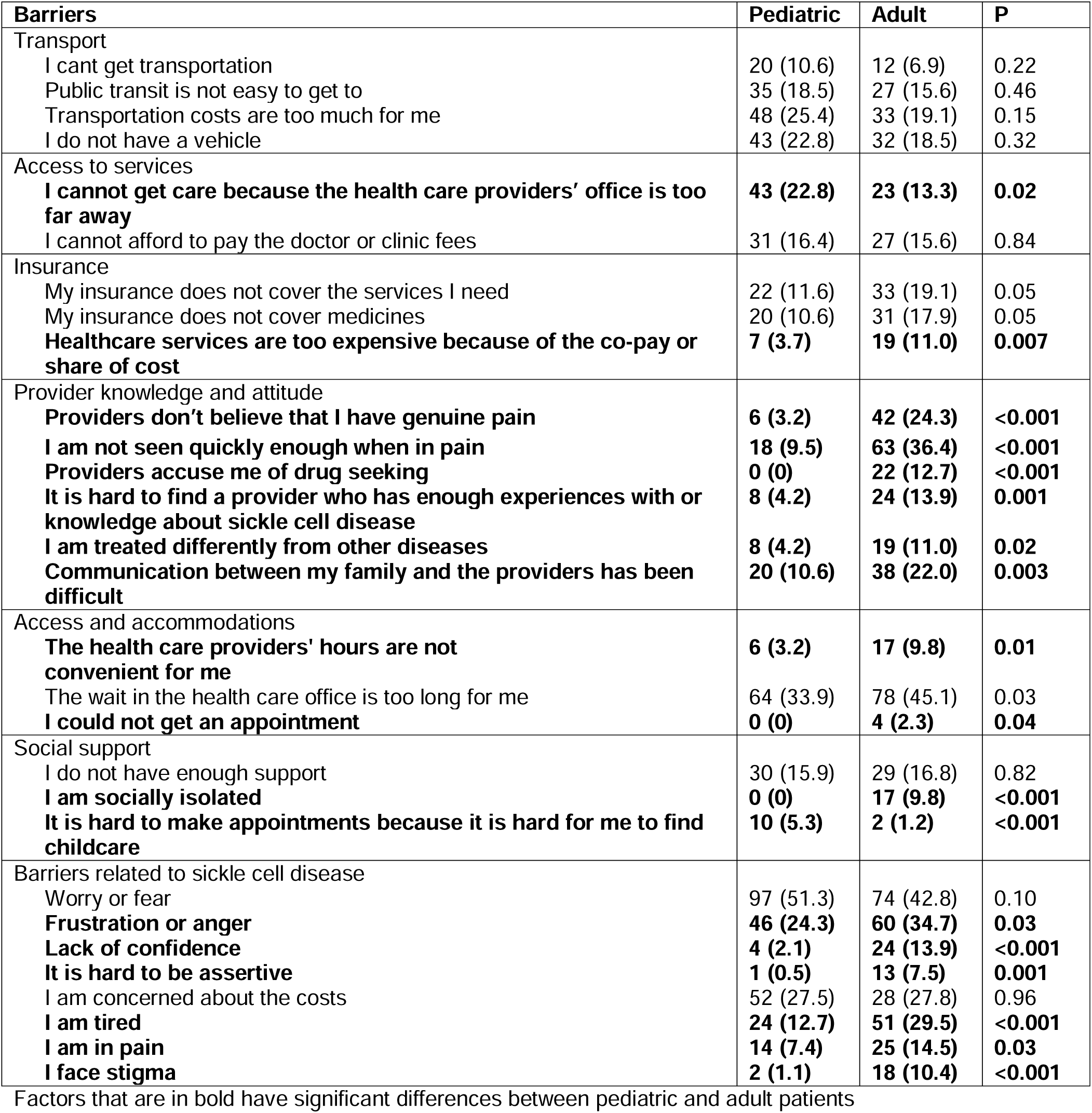
Barriers to Healthcare access reported by Jamaicans with sickle cell disease and their caretakers.

### Hydroxyurea utilization

Overall, 39.4% of children and 30.2% of adults reported initiating hydroxyurea at some point (p=0.18). Nearly two-thirds of adults (62.4%) and parents/caregivers (67.6%) reported having heard of hydroxyurea; 45% adults and 55% of parents/caregivers had been counselled about its use (Figure 1), with no difference between both groups (p>0.05). More children than adults (51% vs. 44%) had been advised to take the drug. Among those patients advised, 77.1% children and 68.4% adults (p=0.179) initiated the drug at some point, and 73.4% of children and 50.0% of adults (p=0.001) are currently taking it. Only 30.2% of the entire adult sample and 40.0% of the entire pediatric sample reported ever taking hydroxyurea. Two-thirds of the cohort reported “always” taking their meds while another third reported “sometimes/not usually” (children: always 67.6% and sometime/not usually 32.4%, vs adult: always 63.2% and sometimes/not usually, 36.8% p=0.64).

**Figure 1:**
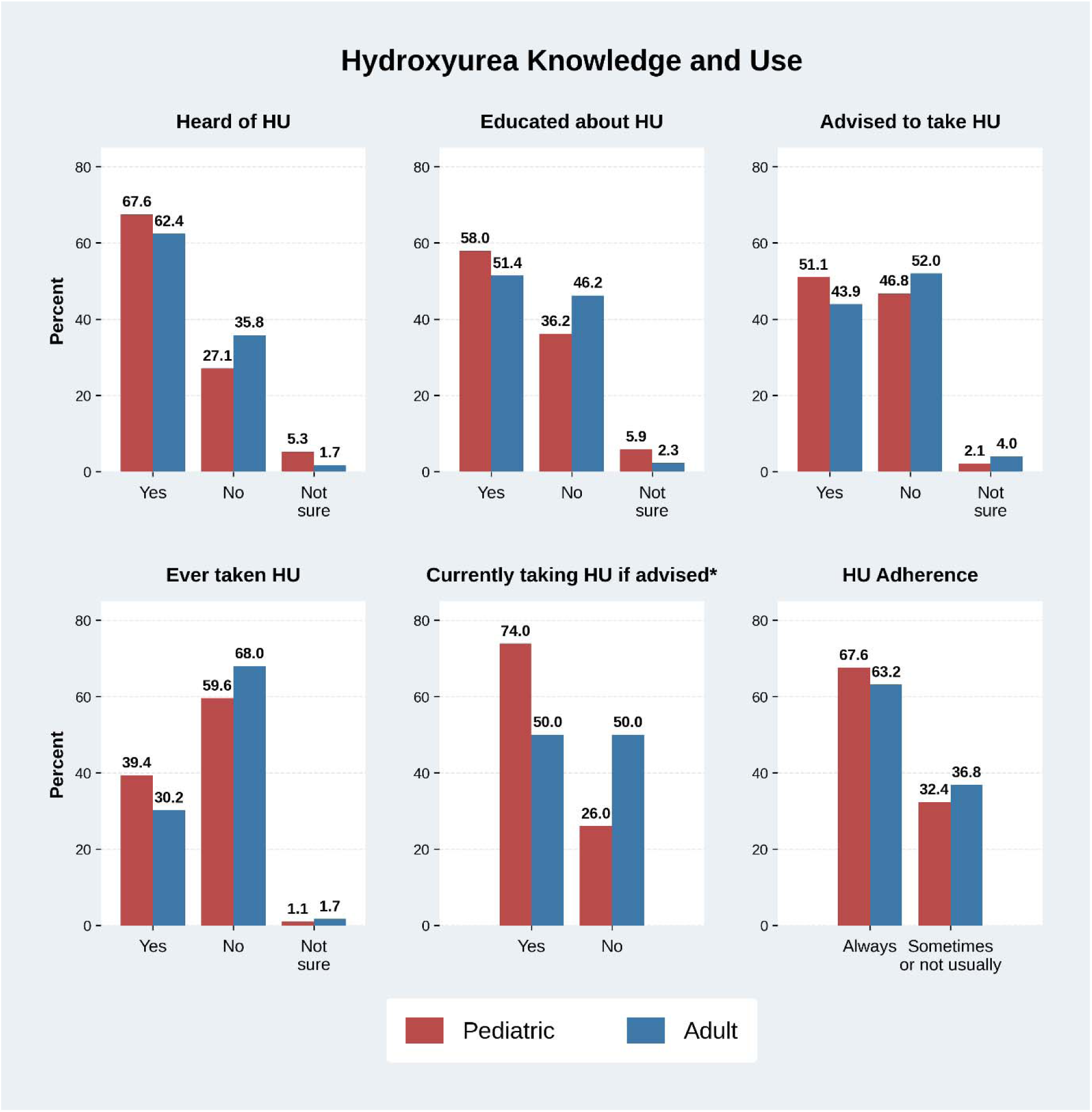
Hydroxyurea Knowledge and Use Reported by Jamaican Patients with Sickle Cell Disease.

### Barriers to Hydroxyurea use

The most frequently reported barrier to using hydroxyurea in both groups was not knowing enough about the medicine, reported by 54.9% of adults and 40.0% of parents/caregivers (Figure 2). Other common barriers in both groups included concerns about side effects (28.3% adults; 25.0% caregivers), dislike of frequent clinic visits (28.9% adults; 24.5% caregivers), and dislike of frequent blood tests (26.7% adults; 25.5% caregivers). A significant number of adults were ‘not interested in taking another medication’ (23.7%) and ‘didn’t like to think about having SCD when feeling well’ (23.8%).

**Figure 2A:**
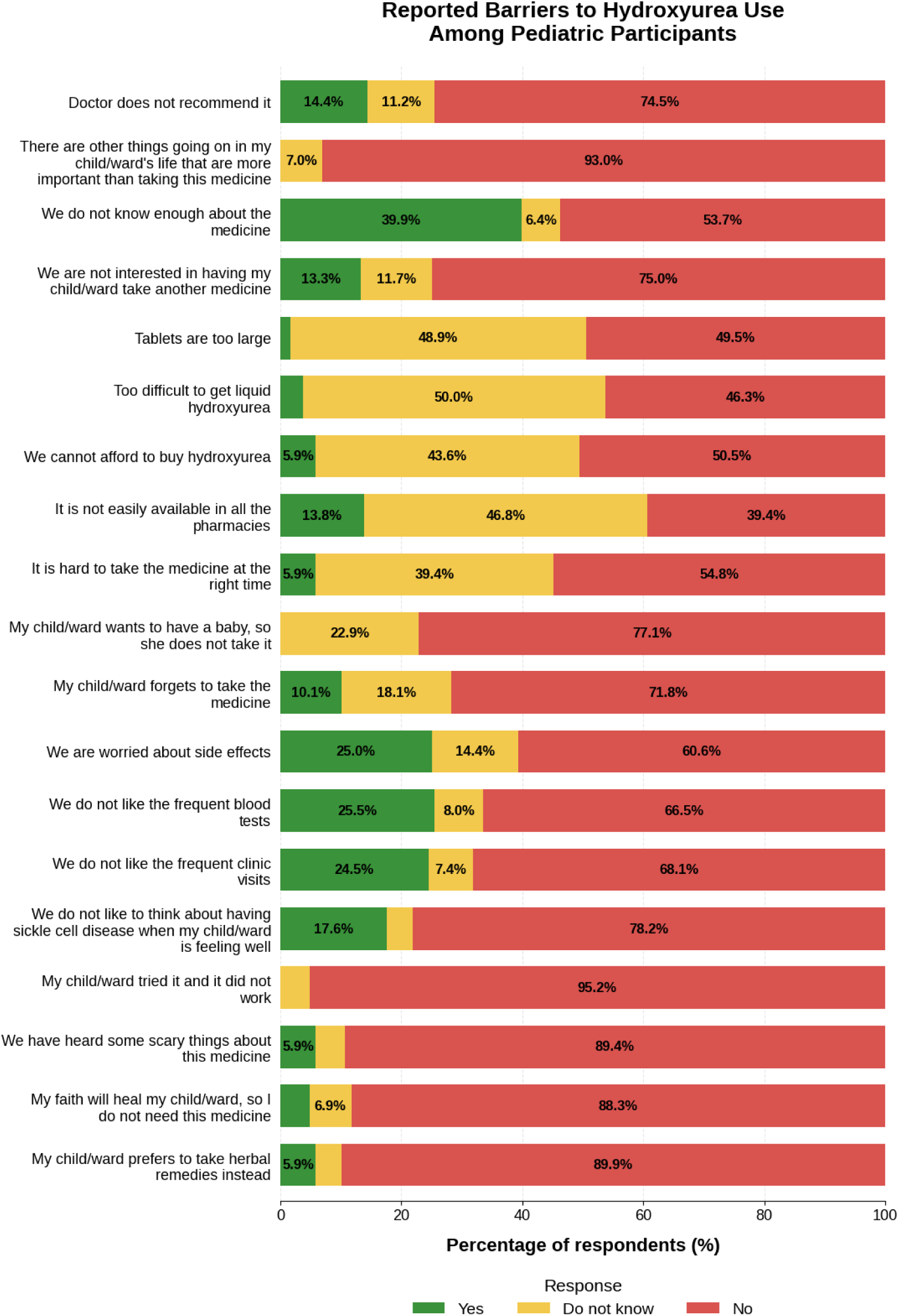
Barriers to hydroxyurea use reported by caretakers of Jamaican pediatric patients with sickle cell disease.

**Figure 2B:**
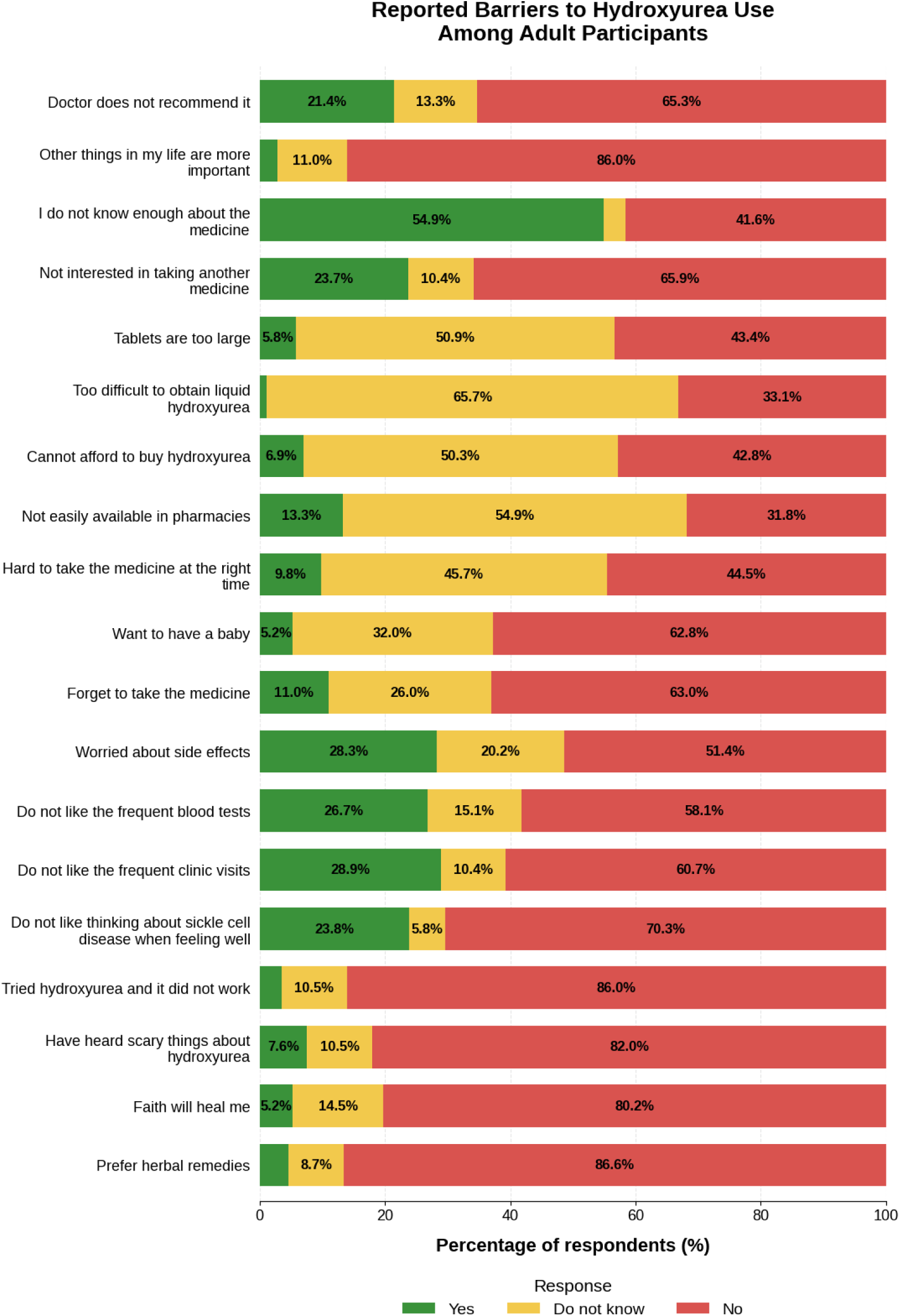
Barriers to hydroxyurea use reported by Jamaican adult patients with sickle cell disease.

### Factors associated with hydroxyurea awareness and education

Adult males reported having heard of and being educated about hydroxyurea less often than adult females (adult male: 47.8% awareness, 41.3% education; female: 67.7% awareness,55.1% education. p<0.05). More adults with tertiary education (77.4%) reported having heard about hydroxyurea compared to those with secondary education (55.7%) (p=0.03). Parents/caregivers who reported hydroyxurea awareness and education had older children (awareness: 7.8±4.6 vs 6.0±5.0 years, p=0.01; education: 8.0±4.6 vs 6.3±4.9 years, p=0.002). No other sociodemographic factors, including rural-urban residence, were associated with hydroxyurea awareness and education.

### Factors associated with hydroxyurea utilization

Among adults, “I don’t know enough about the medicine”, “I am not interested in taking another medicine”, and having no insurance or NHF coverage were negatively associated with hydroxyurea initiation among those advised to take it (Table 4). The same variables were negatively associated with current use of hydroxyurea, although none remained significant after FDR adjustment.

**Table 4:**
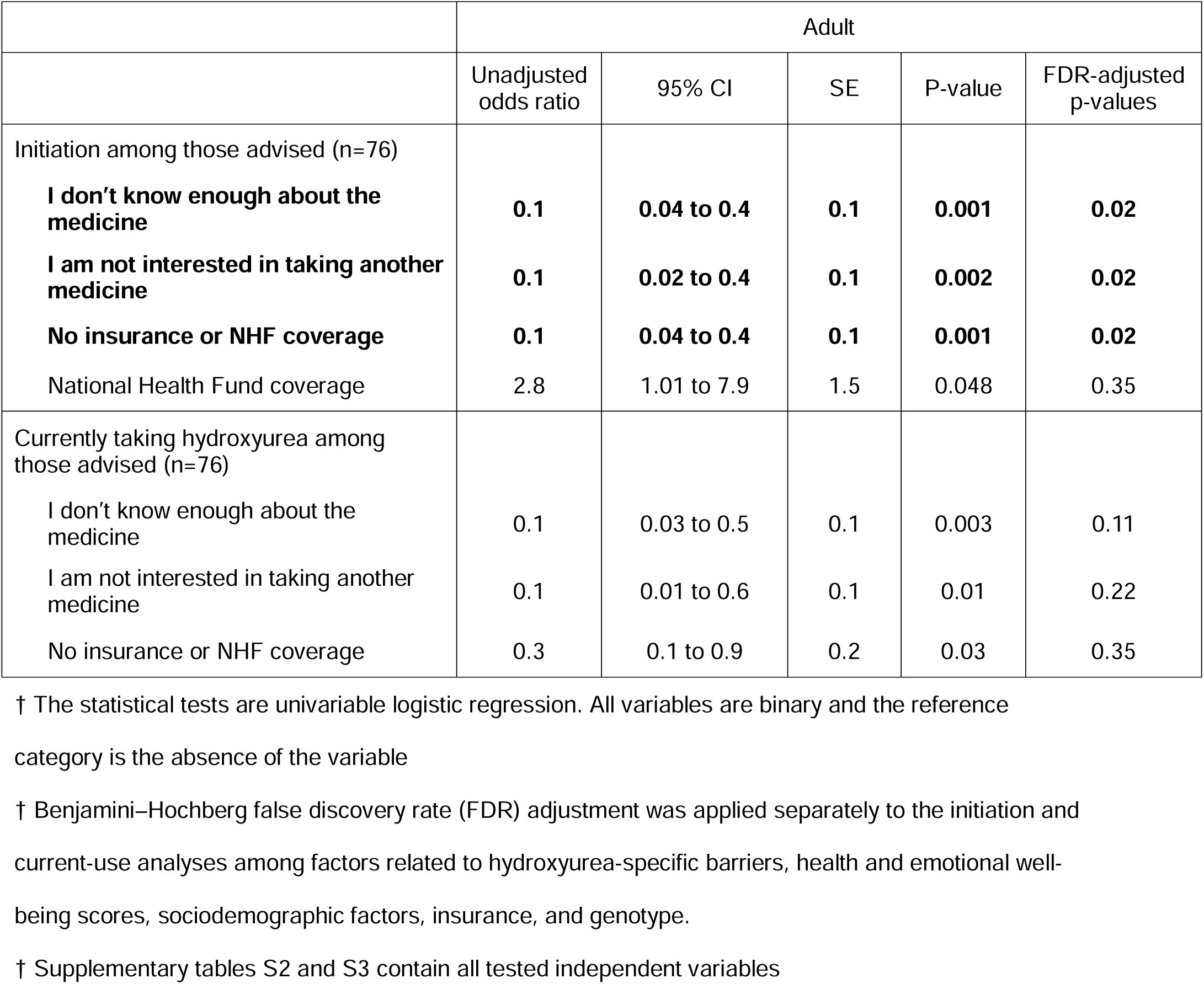
Factors associated with hydroxyurea initiation among adult patients advised to take it.

|  | Adult |  |  |  |  |
| --- | --- | --- | --- | --- | --- |
|  | Unadjusted odds ratio | 95% CI | SE | P-value | FDR-adjusted p-values |
| Initiation among those advised (n=76) |  |  |  |  |  |
| **I don't know enough about the medicine** | **0.1** | **0.04 to 0.4** | **0.1** | **0.001** | **0.02** |
| **I am not interested in taking another medicine** | **0.1** | **0.02 to 0.4** | **0.1** | **0.002** | **0.02** |
| **No insurance or NHF coverage** | **0.1** | **0.04 to 0.4** | **0.1** | **0.001** | **0.02** |
| National Health Fund coverage | 2.8 | 1.01 to 7.9 | 1.5 | 0.048 | 0.35 |
| Currently taking hydroxyurea among those advised (n=76) |  |  |  |  |  |
| I don't know enough about the medicine | 0.1 | 0.03 to 0.5 | 0.1 | 0.003 | 0.11 |
| I am not interested in taking another medicine | 0.1 | 0.01 to 0.6 | 0.1 | 0.01 | 0.22 |
| No insurance or NHF coverage | 0.3 | 0.1 to 0.9 | 0.2 | 0.03 | 0.35 |

† The statistical tests are univariable logistic regression. All variables are binary and the reference category is the absence of the variable
† Benjamini–Hochberg false discovery rate (FDR) adjustment was applied separately to the initiation and current-use analyses among factors related to hydroxyurea-specific barriers, health and emotional well-being scores, sociodemographic factors, insurance, and genotype.
† Supplementary tables S2 and S3 contain all tested independent variables

Among children, hydroxyurea initiation and current use among those advised to take it were higher among older children and those with older parents/caregivers (Table 5). Lower initiation was associated with “we are not interested in having my child/ward take another medicine” and “we are worried about side effects”. In unadjusted analyses, lower current use was associated with “we do not know enough about the medicine”, “we are not interested in having my child/ward take another medicine”, “we are worried about side effects”, and “we do not like to think about SCD when my child/ward is well”.

**Table 5:** Factors associated with hydroxyurea use among paediatric patients advised to take it.

|  | Parent or caretaker |  |  |  |  |
| --- | --- | --- | --- | --- | --- |
|  | Unadjusted odds ratio | 95% CI | SE | P-value | FDR-adjusted p-values |
| Initiation among those advised (n=96) |  |  |  |  |  |
| Age of child | 1.1 | 1.001 to 1.2 | 0.1 | 0.048 | 0.37 |
| Age of parent | 1.1 | 1.02 to 1.2 | 0.04 | 0.01 | 0.13 |
| **We are not interested in having my child/ward take another medicine** | **0.1** | **0.01 to 0.2** | **0.04** | **<0.001** | **<0.001** |
| **We are worried about side effects** | **0.2** | **0.1 to 0.5** | **0.1** | **0.002** | **0.04** |
| Currently taking hydroxyurea among those advised (n=96) |  |  |  |  |  |
| Age of child | 1.2 | 1.001 to 1.3 | 0.07 | 0.02 | 0.21 |
| Age of parent | 1.1 | 1.02 to 1.1 | 0.03 | 0.01 | 0.13 |
| We do not like to think about SCD when my child/ward is ill | 0.2 | 0.1 to 0.9 | 0.2 | 0.02 | 0.40 |
| We don't know enough about the medicine | 0.2 | 0.1 to 0.9 | 0.2 | 0.03 | 0.21 |
| **We are not interested in having my child/ward take another medicine** | **0.02** | **0.002 to 0.2** | **0.03** | **0.001** | **0.04** |
| **We are worried about side effects** | **0.2** | **0.1 to 0.6** | **0.1** | **0.002** | **0.04** |

† The statistical tests are univariable logistic regressions. All variables except age are binary and the reference category is the absence of the variable
† Benjamini–Hochberg false discovery rate (FDR) adjustment was applied separately to the initiation and current-use analyses among factors related to hydroxyurea-specific barriers, health and emotional well-being scores, sociodemographic factors, insurance, and genotype.
† Supplementary tables S4 and S5 contain all tested independent variables

### Factors associated with adherence

Among adults, lower adherence levels were associated with forgetting to take hydroxyurea and at the right time (27.3% adherent vs 72.7% non-adherent, p=0.003), inadequate social support (28.6% adherent vs 71.4% non-adherent, p=0.04), and the drug being expensive due to co-pay (20.0% adherent vs 80.0% non-adherent, p=0.03). Adherence was lower among male children (adherent: male 56.8% vs female 79.4%, p=0.04) but greater among younger children and those at lower levels of school (adherent: median age 8 IQR(4 to 10) vs non-adherent: median age 11 IQR(8 to 14), p=0.01), as well as those with more educated parents (adherent: primary 25.0% vs secondary 63.8% vs tertiary 85.0%, p=0.04). Forgetting to take the medication was also associated with poor adherence among parents/caregivers (parents/caregivers: 36.4% adherent vs 63.6% non-adherent, p=0.02).

## Discussion

SCD care in low- and middle-income countries is frequently affected by health system and socioeconomic constrictions, including limited technological, diagnostic and therapeutic constraints with resources concentrated within specialized centres [11, 29, 30]. This study aimed to assess the barriers and enablers to healthcare access, including hydroxyurea use, among Jamaicans living with or affected by SCD, including healthcare delivery, financial, insurance, transportation and psychosocial domains. Long wait times, particularly when patients were in pain, emerged as an important barrier. A national survey from the United States also found that patients with SCD waited more than 25% longer in emergency departments compared to patients with other illnesses [31]. Additionally, a study in 2019 among Jamaicans with SCD who were recently discharged from hospital, more than 60% reported wait times for SCD care is too long, though there were no differences between private and public institutions [32]. Furthermore, timely administration of opioids is particularly important during vaso-occlusive crisis treatment, as delays have been associated with poorer outcomes. For example, administration of the second dose of opioids within 30 minutes of the first dose among SCD patients being treated for acute pain in the emergency department has been associated with reduced hospitalizations [33]. These findings reinforce the importance of reducing delays in acute pain management for patients with SCD.

Transportation costs were also frequently reported as a barrier. Transportation challenges may lead to missed appointments and delayed treatment for acute complications. In addition to the direct practical difficulty of physically arriving at healthcare facilities, challenges with transportation-related costs may also be a proxy for low socioeconomic status. A previous study among Jamaicans with SCD found that using public transportation as opposed to a private vehicle was associated with greater perceived unaffordability of healthcare [32]. Worry, fear, anger, and frustration relating to the illness were also frequently reported as barriers to healthcare access. The impact of these mental health symptoms on healthcare access is likely reciprocal, as worsening disease may exacerbate socioeconomic and psychosocial challenges which in turn make it more difficult to access care and contribute to further disease progression. Together, these findings suggest that barriers to healthcare access are multifactorial and include provider-level, socioeconomic and psychosocial challenges.

Multidisciplinary interventions at both policy and health system-levels are therefore needed to address these barriers. Emergency department protocols aimed at reducing the time to analgesic administration, particularly opioids, have been designed and successfully implemented in various settings [34–37]. This rapid analgesia approach is also a conditional recommendation of the American Society of Hematology 2020 guidelines for sickle cell disease: management of acute and chronic pain [38]. Access to mental health support, and consequently the management of depression and anxiety, may be improved by integrating psychological and psychiatric services into routine care [39]. Jamaica has adopted several strategies to address transportation and access challenges including providing a bus that transports patients to the specialized SCD center, decentralizing care by training doctors across the island in SCD management, and providing telehealth visits [40]. However, further efforts, supported by additional funding, are needed, as well as improved access to essential therapies.

Hydroxyurea has been the only widely available disease-modifying therapy for SCD for almost 30 years [41]. Although evidence from randomized controlled trials has established its efficacy and utility [13, 42], uptake remains low worldwide [43], with uptake as low as 30% or less among eligible patients reported in the United States [18, 44]. Jamaica boasts a comprehensive SCD care centre of excellence (SCU) since the last 4 decades and has used hydroxyurea in its clinical care protocols for over two decades. However, hydroxyurea knowledge, uptake and barriers to its use have not been previously well characterized. In this study, approximately two-thirds of the participants reported hearing about and receiving counselling or education on hydroxyurea. These rates are lower than the reported awareness and counselling of more than 80% reported in developing African countries [45, 46]. Overall, about one-third of the participants reported initiating hydroxyurea at some point in their lives. However, among participants who reported being advised to use hydroxyurea, more than two-thirds had initiated treatment. Notably, these rates are not directly comparable with population-level estimates of hydroxyurea uptake among clinically eligible patients, as the denominators differ.

In the United States, clinical guidelines have broadened the criteria for eligibility to include early initiation among all patients with SS disease above nine months of age, whereas Jamaica still focuses mostly on criteria related to severity of disease [14]. Additionally, the overall initiation estimates observed in this study should be interpreted cautiously, as a substantial proportion of the participants were recruited from a specialized SCD center where access and knowledge are expected to be greater. By comparison, the multicenter Sickle Cell Disease Implementation Consortium (SCDIC) Needs Assessment study found that 87% of the patient participants had taken hydroxyurea at some point in their lives [47]. Nevertheless, significant efforts are needed to improve hydroxyurea initiation, continuation, and adherence among eligible patients. Given the low awareness and that insufficient knowledge of the drug was associated with lower initiation and current use, greater emphasis should be placed on strengthening hydroxyurea education.

We also aimed to assess various sociodemographic and clinical factors associated with hydroxyurea use, as well as assess barriers to its use. We found that hydroxurea awareness and education were generally more frequently reported among higher educated parents, and parents/caregivers who perceived their knowledge of hydroxyurea to be adequate were more likely to have children who had ever used hydroxyurea. Children with older parents/caregivers were also more likely to initiate and adhere to hydroxyurea therapy. Higher parental education may be associated with greater exposure to more SCD counselling and education, as well as greater capacity to understand and act on the information. Older parents/caregivers may also have gained more experience navigating SCD care which may have contributed to their ability to make informed healthcare decisions. Numerous studies have found that greater parental knowledge is associated with greater medication adherence among SCD patients, including hydroxyurea and deferoxamine [48–50]. Persons with lower health literacy have been shown to be less likely to trust information from specialist doctors and more likely to trust social media, television and friends which may result in lower quality information (28). Educational strategies to improve hydroxyurea use will need to target these persons with lower health literacy and those who may face greater difficulty accessing, understanding, or acting on health information. Published interventions among persons with low literacy have successfully utilized video coaching, audio recordings and brochures written at a reading level of grade nine or lower [51], while the use of community health workers (CHW) may help parents/caregivers and patients navigate the care pathway and improve adherence [52]. These strategies should also account for important differences in hydroxyurea knowledge, use and adherence.

Notably, the proportion of participants who were still taking hydroxyurea differed significantly between children and adults, with more adults reporting that they had discontinued the drug. Adult patients with SCD often report having less support, both financially and from healthcare providers, and suffer from new challenges such as stigmatization, compared with children with SCD [53]. These changes after the ‘transition period’ may explain some of the differences in the variation of hydroxyurea use and adherence. There were also important observed sex differences, as adult females reported higher hydroxyurea awareness and education, while female children were more likely to be adherent. Previous studies have identified important multifactorial sex differences in medication adherence and related health-literacy and self-management, with females tending to adopt more pre-emptive self-care and healthcare engagement, as opposed to males who tend to rely more on support from others while being less likely to seek assistance [54]. The findings in this study highlight the need to develop strategies that more effectively engage males and are tailored to sex-specific differences in health-seeking behaviour, treatment education and self-management support. Successful strategies previously employed focus on ensuring the communication is adapted specifically to reach men, intentionally organizing and structuring the treatment around men’s needs, and establishing a strong relationship between men and the healthcare providers [55].

We found that insurance coverage and NHF enrolment were associated with greater uptake and continuation of hydroxyurea use, while poor adherence was linked with not having adequate social support and the drug being expensive due to co-pay. In a previous study among Jamaican patients recently discharged from hospital, only 16.5% reported having health insurance and 39.8% owned NHF drug-subsidy cards [32]. Although the availability of insurance and whether it covers hydroxurea varies, the findings of this study suggest that the NHF, which subsidizes the cost of hydroxyurea, remains substantially underutilized in this population. Efforts should prioritize improving patient education and awareness, alongside facilitating NHF enrolment and access. Beyond these financial barriers to hydroxyurea use, previous studies have distinguished intentional barriers, such as fears about side effects, and non-intentional barriers, such as forgetting to take the medication [56]. Notably in this study, concerns about side effects were associated with discontinuation and forgetting to take the medication was the main reason identified for non-adherence. These findings highlight the importance of counselling patients about anticipated side effects and how they can be managed. Targeted efforts to improve adherence, including telehealth, CHWs, and mHealth tools that provide medication reminders, may also be considered [52, 57, 58].

## Conclusion

The study has identified multiple barriers to accessing care for SCD. Greater support is needed to address mental health and transportation challenges, alongside the implementation of protocols aimed at reducing wait times. Hydroxyurea awareness and education are inadequate and both initiation and continued use remain suboptimal, highlighting the need for improved education strategies and NHF enrolment. Strategies should be tailored to account for important differences in how males, parents and patients of different age groups, and those with lower educational attainment are engaged.

## Limitations

A substantial proportion of the sample was derived from a specialized SCD center, as opposed to regional health districts, and this may overestimate the awareness, education and use of hydroxyurea.

## Acknowledgements

Data collection instruments were used/modified with permission from those developed under the Sickle Cell Disease Implementation Consortium supported by cooperative agreements from the National Heart, Lung, and Blood Institute and the National Institute on Minority Health and Health Disparities (Bethesda, MD).

## Author contributions

ZR: Data Curation, Formal Analysis, Writing – Original Draft Preparation, Writing – Review & Editing

SM: Conceptualization, Funding Acquisition, Methodology, Project Administration, Writing – Review & Editing

OB: Conceptualization, Investigation, Writing – Review & Editing

VC: Conceptualization, Investigation, Methodology, Project Administration, Writing – Review & Editing

JKM: Conceptualization, Methodology, Writing – Review & Editing

JB: Conceptualization, Investigation, Writing – Review & Editing

JH: Conceptualization, Investigation, Project Administration, Writing – Review & Editing

JRP: Conceptualization, Investigation, Writing – Review & Editing

MA: Conceptualization, Formal Analysis, Data Curation, Funding Acquisition, Methodology, Project Administration, Writing – Original Draft Preparation, Writing – Review & Editing

## Data availability

The dataset is not publicly available due to ethical and privacy considerations. Data may be made available upon reasonable request and subject to appropriate approvals.

