## Supplemental Table 1 for "Sickle Cell Disease Care in Jamaica: Enablers and Barriers to Healthcare Access and Hydroxyurea Utilization"

**S1 Table: Barriers to Healthcare access reported by Jamaicans with sickle cell disease and their caretakers**

| **Barriers** | **Pediatric** | **Adult** | **P** |
| --- | --- | --- | --- |
| Transport |  |  |  |
| I cant get transportation | 20 (10.6) | 12 (6.9) | 0.22 |
| Public transit is not easy to get to | 35 (18.5) | 27 (15.6) | 0.46 |
| Transportation costs are too much for me | 48 (25.4) | 33 (19.1) | 0.15 |
| I do not have a vehicle | 43 (22.8) | 32 (18.5) | 0.32 |
| I do not have access to a vehicle | 27 (14.3) | 18 (10.4) | 0.26 |
| Access to services |  |  |  |
| I do not know where to get care | 1 (0.5) | 2 (1.2) | 0.51 |
| **I cannot get care because the health care providers’ office is too far away** | **43 (22.8)** | **23 (13.3)** | **0.02** |
| I cannot afford to pay the doctor or clinic fees | 31 (16.4) | 27 (15.6) | 0.84 |
| I do not want to go out because of COVID-19 | 7 (3.7) | 2 (1.2) | 0.12 |
| Insurance |  |  |  |
| My insurance does not cover the services I need | 22 (11.6) | 33 (19.1) | 0.05 |
| My insurance does not cover medicines | 20 (10.6) | 31 (17.9) | 0.05 |
| **Healthcare services are too expensive because of the co-pay or share of cost** | **7 (3.7)** | **19 (11.0)** | **0.007** |
| It takes too long to get approval for the care that I need | 0 (0) | 1 (0.6) | 0.30 |
| Getting reimbursements for some treatments or services is hard | 2 (1.1) | 6 (3.5) | 0.12 |
| Provider knowledge and attitude |  |  |  |
| **Providers don’t believe that I have genuine pain** | **6 (3.2)** | **42 (24.3)** | **<0.001** |
| **I am not seen quickly enough when in pain** | **18 (9.5)** | **63 (36.4)** | **<0.001** |
| **Providers accuse me of drug seeking** | **0 (0)** | **22 (12.7)** | **<0.001** |
| Providers let me know that they do not appreciate how knowledgeable I am about the disease | 6 (3.2) | 8 (4.6) | 0.48 |
| **It is hard to find a provider who has enough experiences with or knowledge about sickle cell disease** | **8 (4.2)** | **24 (13.9)** | **0.001** |
| **I am treated differently from other diseases** | **8 (4.2)** | **19 (11.0)** | **0.02** |
| **Communication between my family and the providers has been difficult** | **20 (10.6)** | **38 (22.0)** | **0.003** |
| Access and accommodations |  |  |  |
| Places for me to go to learn how to stay well are not close by or easy to get to | 14 (7.4) | 10 (5.8) | 0.53 |
| **The health care providers' hours are not**  **convenient for me** | **6 (3.2)** | **17 (9.8)** | **0.01** |
| The wait in the health care office is too long for me | 64 (33.9) | 78 (45.1) | 0.03 |
| The paperwork I have to fill out is too much | 2 (1.1) | 1 (0.6) | 0.62 |
| **I could not get an appointment** | **0 (0)** | **4 (2.3)** | **0.04** |
| Social support |  |  |  |
| I do not have enough support | 30 (15.9) | 29 (16.8) | 0.82 |
| The people who take care of me or give me support are burned out | 7 (3.7) | 17 (9.8) | 0.02 |
| I am burned out by taking care of others or giving support to them | 9 (4.8) | 5 (2.9) | 0.36 |
| I need help with daily chores | 9 (4.8) | 12 (6.9) | 0.38 |
| **I am socially isolated** | **0 (0)** | **17 (9.8)** | **<0.001** |
| There are other things going on in my family that  are more important than my health care | 10 (5.3) | 3 (1.7) | 0.07 |
| **It is hard to make appointments because it is hard for me to find childcare** | **100 (5.3)** | **2 (1.2)** | **<0.001** |
| Barriers for individuals |  |  |  |
| I don't really know what to do to stay healthy | 19 (10.1) | 10 (5.8) | 0.14 |
| I don't know enough about the sickle cell disease care that I need | 26 (13.8) | 21 (12.1) | 0.65 |
| I don't understand the system or find it too hard to work through | 4 (2.1) | 7 (4.1) | 0.29 |
| It is hard to follow up on care | 8 (4.2) | 14 (8.1) | 0.13 |
| I miss appointments because of memory problems | 9 (4.8) | 11 (6.4) | 0.51 |
| The medical instructions are hard to follow | 2 (1.1) | 2 (1.1) | 0.93 |
| Staff are hard to talk to | 9 (4.8) | 7 (4.1) | 0.74 |
| Staff are hard to understand | 0 (0) | 4 (2.3) | 0.04 |
| Barriers related to sickle cell disease |  |  |  |
| Worry or fear | 97 (51.3) | 74 (42.8) | 0.10 |
| **Frustration or anger** | **46 (24.3)** | **60 (34.7)** | **0.03** |
| **Lack of confidence** | **4 (2.1)** | **24 (13.9)** | **<0.001** |
| **It is hard to be assertive** | **1 (0.5)** | **13 (7.5)** | **0.001** |
| It is embarrassing | 3 (1.6) | 13 (7.5) | 0.006 |
| I am concerned about the costs | 52 (27.5) | 28 (27.8) | 0.96 |
| **I am tired** | **24 (12.7)** | **51 (29.5)** | **<0.001** |
| **I am in pain** | **14 (7.4)** | **25 (14.5)** | **0.03** |
| **I face stigma** | **2 (1.1)** | **18 (10.4)** | **<0.001** |
