## Supplemental Table 3 for "Sickle Cell Disease Care in Jamaica: Enablers and Barriers to Healthcare Access and Hydroxyurea Utilization"

**S3 Table: Factors associated with current use of hydroxyurea among Jamaican adults with sickle cell disease advised to take it**

| **Factor** | **Odds ratio** | **95% CI lower** | **95% CI upper** | **p-value** | **FDR-adjusted p-value** |
| --- | --- | --- | --- | --- | --- |
| **Hydroxyurea-specific barrier** |  |  |  |  |  |
| Doctor does not recommend hydroxyurea* | 0.1 | 0.02 | 1.2 | 0.08 | 0.35 |
| Insufficient knowledge about hydroxyurea* | 0.1 | 0.03 | 0.5 | 0.003 | 0.11 |
| Not interested in taking another medication* | 0.1 | 0.01 | 0.6 | 0.01 | 0.22 |
| Hydroxyurea tablets are too large* | 2.3 | 0.4 | 12.6 | 0.32 | 0.63 |
| Too difficult to obtain liquid hydroxyurea* | 1.2 | 0.1 | 19.8 | 0.92 | 0.95 |
| Cannot afford hydroxyurea* | 1.7 | 0.4 | 7.4 | 0.49 | 0.73 |
| Hydroxyurea is not easily available in pharmacies* | 2.2 | 0.7 | 6.7 | 0.17 | 0.46 |
| Difficulty taking hydroxyurea at the right time* | 2.9 | 0.7 | 11.6 | 0.14 | 0.40 |
| Fertility or pregnancy plans* | 0.3 | 0.03 | 2.7 | 0.26 | 0.55 |
| Forgets to take hydroxyurea* | 3.8 | 0.95 | 15.1 | 0.06 | 0.35 |
| Concern about side effects* | 0.9 | 0.3 | 2.2 | 0.76 | 0.85 |
| Dislikes frequent blood tests* | 0.4 | 0.1 | 1.1 | 0.07 | 0.35 |
| Dislikes frequent clinic visits* | 0.6 | 0.2 | 1.5 | 0.27 | 0.55 |
| Does not like thinking about SCD when feeling well* | 1.1 | 0.4 | 3.2 | 0.82 | 0.89 |
| Has heard scary things about hydroxyurea* | 0.5 | 0.1 | 2.1 | 0.36 | 0.67 |
| Faith will heal SCD so hydroxyurea is not needed* | 0.4 | 0.04 | 5.1 | 0.51 | 0.73 |
| Prefers herbal remedies instead* | 1.0 | 0.2 | 5.3 | 1.00 | 1.00 |
| Reports no hydroxyurea barriers* | 2.8 | 0.8 | 10.0 | 0.12 | 0.40 |
| **Health and emotional well-being** |  |  |  |  |  |
| Current health score | 1.1 | 0.7 | 1.8 | 0.64 | 0.83 |
| SCD emotional well-being score | 0.9 | 0.7 | 1.2 | 0.49 | 0.73 |
| **Sociodemographics** |  |  |  |  |  |
| Sex* | 0.7 | 0.3 | 2.2 | 0.59 | 0.81 |
| Residence* | 2.4 | 0.9 | 6.1 | 0.07 | 0.35 |
| Educational attainment* | 0.4 | 0.2 | 1.1 | 0.07 | 0.35 |
| Married (reference: never married) | 0.2 | 0.02 | 1.5 | 0.11 | 0.40 |
| Common-law (reference: never married) | 0.4 | 0.1 | 1.8 | 0.25 | 0.55 |
| Visiting relationship (reference: never married) | 1.4 | 0.3 | 6.4 | 0.68 | 0.83 |
| Monthly household income $27,001–47,999 (reference: ≤$27,000) | 3.0 | 0.2 | 42.6 | 0.42 | 0.70 |
| Monthly household income $48,000–99,999 (reference: ≤$27,000) | 0.4 | 0.04 | 3.6 | 0.40 | 0.70 |
| Monthly household income $100,000–249,000 (reference: ≤$27,000) | 2.0 | 0.3 | 14.8 | 0.50 | 0.73 |
| Monthly household income >$250,000 (reference: ≤$27,000) | 0.9 | 0.1 | 5.6 | 0.89 | 0.94 |
| Monthly household income Don't know/not sure (reference: ≤$27,000) | 0.7 | 0.1 | 3.8 | 0.66 | 0.83 |
| Monthly household income No response (reference: ≤$27,000) | 0.2 | 0.03 | 1.6 | 0.14 | 0.40 |
| **Insurance** |  |  |  |  |  |
| Self-purchased insurance* | 1.3 | 0.3 | 5.2 | 0.72 | 0.84 |
| Employer-provided insurance* | 2.0 | 0.7 | 5.5 | 0.20 | 0.50 |
| NHF coverage* | 2.4 | 0.9 | 6.8 | 0.08 | 0.35 |
| Neither NHF nor insurance* | 0.3 | 0.1 | 0.9 | 0.03 | 0.35 |
| **Genotype** |  |  |  |  |  |
| Severe genotype* | 1.4 | 0.3 | 6.6 | 0.69 | 0.83 |

*Binary variables (the reference category is the absence of the variable)

† The statistical tests are univariable logistic regressions.
