## Supplemental Table 4 for "Sickle Cell Disease Care in Jamaica: Enablers and Barriers to Healthcare Access and Hydroxyurea Utilization"

**S4 Table: Factors associated with the initiation of hydroxyurea among parents/caregivers of children with sickle cell disease advised to take it**

| **Factor** | **Odds ratio** | **95% CI lower** | **95% CI upper** | **p-value** | **FDR-adjusted p-value** |
| --- | --- | --- | --- | --- | --- |
| **Hydroxyurea-specific barrier** |  |  |  |  |  |
| Doctor does not recommend hydroxyurea* | 0.3 | 0.1 | 1.5 | 0.13 | 0.49 |
| Insufficient knowledge about hydroxyurea* | 0.3 | 0.1 | 1.1 | 0.07 | 0.40 |
| Not interested in child taking another medication* | 0.05 | 0.01 | 0.2 | <0.001 | <0.001 |
| Cannot afford hydroxyurea* | 0.9 | 0.2 | 4.8 | 0.92 | 0.92 |
| Hydroxyurea is not easily available in pharmacies* | 3.2 | 0.7 | 15.3 | 0.15 | 0.51 |
| Difficulty giving hydroxyurea at the right time* | 1.3 | 0.1 | 11.2 | 0.84 | 0.92 |
| Concern about side effects* | 0.2 | 0.1 | 0.5 | 0.002 | 0.04 |
| Dislikes frequent blood tests* | 0.6 | 0.2 | 1.6 | 0.30 | 0.65 |
| Dislikes frequent clinic visits* | 0.6 | 0.2 | 1.7 | 0.34 | 0.65 |
| Does not like thinking about SCD when child is well* | 0.3 | 0.1 | 1.1 | 0.07 | 0.40 |
| Has heard scary things about hydroxyurea* | 0.5 | 0.1 | 2.1 | 0.32 | 0.65 |
| Prefers herbal remedies instead* | 0.3 | 0.04 | 2.1 | 0.21 | 0.56 |
| **Health and emotional well-being** |  |  |  |  |  |
| Child emotional well-being score | 1.2 | 0.9 | 1.4 | 0.20 | 0.56 |
| Child current health score | 0.9 | 0.5 | 1.4 | 0.57 | 0.85 |
| **Sociodemographics** |  |  |  |  |  |
| Child sex* | 0.7 | 0.3 | 1.8 | 0.46 | 0.79 |
| Child in kindergarten (reference: not yet attended school) | 2.0 | 0.6 | 6.7 | 0.26 | 0.62 |
| Child in primary school (reference: not yet attended school) | 3.8 | 0.8 | 18.8 | 0.10 | 0.49 |
| Parent/caregiver educational attainment* | 0.4 | 0.1 | 1.5 | 0.17 | 0.54 |
| Parent/caregiver married (reference: never married) | 5.1 | 1.01 | 25.3 | 0.048 | 0.37 |
| Parent/caregiver separated/divorced (reference: never married) | 1.8 | 0.2 | 17.6 | 0.63 | 0.86 |
| Parent/caregiver common-law relationship (reference: never married) | 1.3 | 0.4 | 3.8 | 0.69 | 0.87 |
| Parent/caregiver visiting relationship (reference: never married) | 0.9 | 0.1 | 10.8 | 0.92 | 0.92 |
| Monthly household income $27,001–47,999 (reference: ≤$27,000) | 0.5 | 0.04 | 5.2 | 0.52 | 0.85 |
| Monthly household income $48,000–99,999 (reference: ≤$27,000) | 0.6 | 0.04 | 8.7 | 0.71 | 0.87 |
| Monthly household income $100,000–249,000 (reference: ≤$27,000) | 0.4 | 0.04 | 4.2 | 0.45 | 0.79 |
| Monthly household income >$250,000 (reference: ≤$27,000) | 2.2 | 0.1 | 42.7 | 0.60 | 0.85 |
| Monthly household income Don't know/not sure (reference: ≤$27,000) | 0.8 | 0.1 | 8.4 | 0.82 | 0.92 |
| Monthly household income No response (reference: ≤$27,000) | 0.8 | 0.1 | 8.9 | 0.86 | 0.92 |
| Parent/caregiver unemployed and looking (reference: full-time) | 1.6 | 0.3 | 8.0 | 0.59 | 0.85 |
| Parent/caregiver unemployed and not looking (reference: full-time) | 0.9 | 0.2 | 4.9 | 0.87 | 0.92 |
| Parent/caregiver student (reference: full-time) | 0.7 | 0.1 | 8.1 | 0.77 | 0.92 |
| Crowding index | 0.9 | 0.6 | 1.4 | 0.68 | 0.87 |
| Parent/caregiver age | 1.1 | 1.02 | 1.2 | 0.01 | 0.13 |
| Child age | 1.1 | 1.001 | 1.3 | 0.048 | 0.37 |
| **Insurance** |  |  |  |  |  |
| Employer-provided insurance* | 1.3 | 0.5 | 3.6 | 0.61 | 0.85 |
| NHF coverage* | 1.8 | 0.7 | 4.9 | 0.22 | 0.56 |
| Neither NHF nor insurance* | 0.6 | 0.2 | 1.6 | 0.33 | 0.65 |
| **Genotype** |  |  |  |  |  |
| Severe genotype* | 3.7 | 0.7 | 20.0 | 0.12 | 0.49 |

*Binary variables (the reference category is the absence of the variable)

† The statistical tests are univariable logistic regressions.
